# The association between peripheral neuropathy and daily-life gait quality characteristics in people with diabetes

**DOI:** 10.1101/2024.02.21.24303128

**Authors:** Chantal M Hulshof, Marike van der Leeden, Jaap J van Netten, Maarten Gijssel, Jordi Evers, Sicco A Bus, Mirjam Pijnappels

**Author notes:** Corresponding authors: Chantal M Hulshof, and Jaap J van Netten, Amsterdam UMC location University of Amsterdam, Rehabilitation Medicine, Meibergdreef 9, 1105 AZ, Amsterdam, The Netherlands.

## Abstract

0.

**Background:** Peripheral neuropathy is a common complication of diabetes and increases the risk of falls, possibly through gait (quality) impairments in daily life. Characteristics of gait quality have been associated with peripheral neuropathy in a laboratory setting, but little is known about the more relevant association with gait quality in daily life.

**Research question:** What is the association between peripheral neuropathy and gait quality characteristics in daily life in people with diabetes?

**Methods:** Data from two cross-sectional studies were combined in an exploratory analysis, including a total of 98 participants with diabetes (mean age: 68 (SD 7) years, 32 females), of which 68 with peripheral neuropathy. Participants wore a tri-axial accelerometer for seven consecutive days. Walking episodes ≥5 seconds were identified and analysed to determine various gait quality characteristics. Associations were assessed using linear regression analyses, adjusted for walking speed and other potential confounders.

**Results:** Peripheral neuropathy was significantly associated with a lower walking speed (people with neuropathy: 0.81 vs without neuropathy: 0.88 m/s; β (95% confidence interval (CI)): -0.114 (−0.202 to -0.026)), a lower stride frequency (0.81 vs 0.85 strides/s; β (95% CI): -0.030 (−0.057 to -0.003)), lower gait intensity (i.e. lower root mean square) in vertical direction (1.38 vs 1.63 m/s^2^; β (95% CI): -0.074 (−0.143 to -0.006)), and less gait symmetry (i.e. lower harmonic ratio) in vertical direction (1.82 vs 2.27; β (95% CI): -0.322 (−0.474 to -0.170)). People with peripheral neuropathy had non-significantly poorer gait quality for most of the other 21 gait quality characteristics.

**Significance:** Peripheral neuropathy seems to negatively affect several gait quality characteristics measured in daily life. These results need to be replicated in future studies and may help to develop targeted gait training to improve gait quality and potentially reduce fall risk in people with diabetes and peripheral neuropathy.

## 1. Introduction

Worldwide, the number of people with diabetes mellitus was estimated at 537 million in 2021 [1], and this is expected to rise to 693 million by 2045 [2]. Peripheral neuropathy is a common complication of diabetes, and occurs in up to 50% of people with the disease [3]. Peripheral neuropathy is characterised by a loss of protective sensation and atrophy and dysfunction of the intrinsic and extrinsic foot muscles, leading to foot deformities and biomechanical abnormalities, which increase the risk of foot ulceration [3,4]. In addition, peripheral neuropathy in people with diabetes is a strong predictor of falls [5]. Peripheral neuropathy impedes the important sensory feedback from the feet to maintain stable in posture and gait. Furthermore, neuropathy may lead to a delayed neuromuscular response after initial foot-ground contact during gait, which alters the gait pattern and also contributes to gait impairment [6,7]. Therefore, insights in the gait pattern of people with diabetes and neuropathy could help understanding their increased fall risk and eventually contribute to fall prevention strategies.

From previous studies, various insights in gait patterns of people with diabetes with and without peripheral neuropathy have been gained. A narrative review described that people with diabetes and neuropathy have a slower walking speed, shorter stride length, larger step width and larger gait variability in both temporal and spatial aspects compared to people without diabetes and people with diabetes without peripheral neuropathy [7]. However, aforementioned gait characteristics were all investigated in an optimal, standardised laboratory setting. This context may not be representative of the varying and more challenging conditions people face in daily life [8]. Using advanced technologies, such as wearable sensors, allow to assess gait performance in daily life. For example, inertial sensor data can be used to quantify spatiotemporal characteristics (e.g. walking speed, stride frequency and stride length), as well as gait quality characteristics (e.g. gait stability, symmetry, variability, intensity and smoothness) [9–13]. Such characteristics have been associated with fall risk in healthy older adults [9–11,14–16]. However, whether daily-life gait quality characteristics are affected in people with diabetes and peripheral neuropathy has not yet been established.

Insights into daily-life gait quality of people with diabetes and peripheral neuropathy may help understanding their increased fall risk and could lead to development of targeted interventions to improve gait quality and reduce fall risk in this vulnerable population. Therefore, our aim was to explore the association between the presence of peripheral neuropathy and gait quality characteristics in daily life in people with diabetes.

## 2. Methods

### 2.1. Design and participants: DIALOAD and DWELL-NL

In this cross-sectional exploratory study, we combined data of 116 participants from the DIALOAD (DIAbetic foot LOAD capacity) and DWELL-NL (Diabetes & WELLbeing Netherlands) studies. In both studies, daily-life gait in people with diabetes (with and without peripheral neuropathy) was assessed; the DIALOAD cohort included people with diabetes at high risk of foot ulceration, while the DWELL-NL cohort included people who had diabetes.

In DIALOAD, 52 participants were included. Inclusion criteria for the DIALOAD study were: 18 years and older, diabetes mellitus, loss of protective sensation, ambulatory, and a recently healed foot ulcer (<1 year) or high barefoot plantar pressures (>600 kPa at any region in either foot). Exclusion criteria were: a current foot ulcer, open amputation wound, active Charcot neuro-osteo arthropathy, use of a walking aid for full support, or critical ischemia (toe pressure <30 mmHg). For the current study, we excluded people from the DIALOAD study who were under the age of 55 (n=8) to ensure that the age criterion for inclusion was consistent across studies and to minimise any potential age-related effect on gait quality. DIALOAD is a multi-centre prospective observational cohort study registered in the Netherlands trial register (registration number: NL8839). Participants were recruited between August 2020 and May 2022 in both centres of Amsterdam UMC and podiatry practice Voeten op Texel, in the Netherlands.

In DWELL-NL, 56 participants were included. Inclusion criteria for the DWELL-NL study were: 55 years and older, diabetes mellitus type 2, ambulatory, and cognitive ability to follow instructions and to understand the Dutch or English questionnaires. Exclusion criteria were: psychiatric or memory problems, inability to walk 4 meters, or inability to get up from a chair without help from others. Participants were recruited between February 2017 up to February 2018 using a flyer available at two primary physiotherapy practices in the North-East of the Netherlands: Move2be and BuurtFysio.

For both studies, written informed consent was obtained from all participants prior to inclusion. All study procedures were in accordance with the Declaration of Helsinki. The requirement for ethical review of the study was waived under the Medical Research Involving Human Subjects Act in the Netherlands by the accredited medical ethics committees of Amsterdam UMC for DIALOAD (registration number: W19_429#19.495) and of Brabant for DWELL-NL (registration number: NL62544.028.17).

### 2.2. Measurements

#### 2.2.1. Peripheral neuropathy – independent variable

Peripheral neuropathy was defined as the presence of loss of protective sensation in the foot. In DIALOAD, loss of protective sensation was assessed using a 10-gram monofilament and tuning fork according to international guidelines [17]. In DWELL-NL, information about the presence of peripheral neuropathy was obtained from the electronic health record system of the general practitioner or medical specialist, and was assessed similarly as in DIALOAD. The presence of peripheral neuropathy was the independent variable in the analysis of the current study.

#### 2.2.2. Gait quality characteristics – dependent variables

In both studies, participants wore a tri-axial accelerometer measuring at a frequency of 100 Hz and at an amplitude range of -6g to +6g (MoveMonitor, McRoberts, The Hague, The Netherlands) at their back at vertebrae level L5 during seven consecutive days immediately following the study visit. The instruction for participants was to wear the accelerometer at all times (including when asleep), except during activities involving water, such as showering and swimming, because the device is not water resistant.

Gait quality characteristics can be reliably estimated from raw acceleration data [9,10]. First, the manufacturer’s algorithm was used to identify walking episodes from the raw accelerometer data. All walking episodes with a minimum duration of 5 seconds were merged and cut into 5-second windows to avoid bias by different lengths of data series [10]. A minimum of 50 walking episodes per participant were required to reliably calculate median values of gait quality characteristics of all 5-second windows [9]. Then, the following gait quality characteristics were derived from the accelerometer data and explained and justified in Table 1: walking speed, stride length, stride length variability, stride frequency, stride regularity, gait complexity (i.e. sample entropy), gait intensity (i.e. root mean square), gait smoothness (i.e. index of harmonicity), gait symmetry (i.e. harmonic ratio), gait consistency (i.e. power at step frequency), gait stability (i.e. Lyapunov estimate and Lyapunov per stride) and a gait quality composite score. Walking speed was estimated based on the inverted pendulum model [18], with leg length (required for this model) estimated as 53% of body height [19]. To estimate gait quality characteristics from the raw acceleration data, we used the Open Access Gait Analysis Toolbox version 3.3 (https://github.com/VU-HMS/Gait-Analysis) provided by the Vrije Universiteit Amsterdam. The Gait Analysis Toolbox was developed to identify fall risk predictors from daily life accelerometry data [9,10].

**Table 1.** Technical and clinical explanation of gait quality characteristics based on daily-life accelerometry data.

| Gait quality characteristic | Technical explanation | Clinical explanation + *interpretation* [12,20] |
| --- | --- | --- |
| **Walking speed (m/s)** | The product of the stride length and stride frequency (see technical explanation below) [11,18]. | The time required to walk a certain distance.<br>*A faster walking speed is indicative of a better gait quality.* |
| **Stride length (m)** | Based on the inverted pendulum model and derived from the leg length and vertical displacement of the trunk [18]. | The anteroposterior distance between the heel floor contact points of the same foot within a single gait cycle [7].<br>*A longer stride length is indicative of a better gait quality.* |
| **Stride length variability (m)** | The standard deviation of the stride length over multiple strides. | The variability in stride length over strides.<br>*A less variable stride length is indicative of a better gait quality.* |
| **Stride frequency (strides/s)** | The stride frequency was estimated from the median of (half) the modal frequencies [9]. | The number of strides taken during a certain time period.<br>*A higher stride frequency is indicative of a better gait quality.* |
| **Stride regularity <sup>a</sup>** | Combining spatial and temporal gait variability, the summed autocovariance for time-lag $s$ normalised to the summed autocovariance with zero lag [21]. | The consistency of the stride-to-stride pattern, considering both spatial and temporal aspects [21].<br>*Higher values of stride regularity (with a maximum of 1) are indicative of a better gait quality.* |
| **Gait complexity <sup>a</sup>**<br>(i.e. sample entropy) | Expressed as **sample entropy**, which is the negative natural logarithm of the conditional probability for similarity in data sequences [22]. | The automaticity of walking, more automatic gait can be performed with less attention, leading to more complex and unpredictable fluctuations in the gait pattern and hence a more flexible and adaptive system [22].<br>*Higher values of sample entropy are indicative of a better gait quality.* |
| **Gait intensity (m/s<sup>2</sup>) <sup>a</sup>**<br>(i.e. root mean square) | Expressed as the **root mean square** of the signal, which is the magnitude of an acceleration signal and calculated by the square root of the mean of the squares [11]. | This reflects the amount of trunk movement during walking within each walking episode [23].<br>*Higher values of the root mean square are indicative of a better gait quality.* |
| **Gait smoothness <sup>a</sup>**<br>(i.e. index of harmonicity) | Expressed as **index of harmonicity**, which was computed by using Fourier transformation and quantifies the degree of periodicity [24]. | The regularity or smoothness of a walking pattern, a value of 1 indicates a perfectly smooth gait [24].<br>*Higher values of the index of harmonicity (with a maximum of 1) are indicative of a better gait quality.* |
| **Gait symmetry <sup>a</sup>**<br>(i.e. harmonic ratio) | Expressed as **harmonic ratio**, which was computed by using Fourier transformation and quantifies the strength of harmonics relative to the fundamental frequency [25,26]. | The symmetry between the right and left steps during walking [26].<br>*Higher values of harmonic ratio are indicative of a better gait quality.* |
| **Gait consistency (psd) <sup>a</sup>**<br>(i.e. power at step frequency) | Expressed as **power at step frequency**, which is the amplitude of the dominant power in the frequency domain [14]. | The consistency or repeatability of a walking pattern within a walking episode [23].<br>*Higher values of the power at step frequency are indicative of a better gait quality.* |
| **Gait stability**<br>(i.e. Lyapunov estimate and Lyapunov per stride) | Expressed as the maximal **Lyapunov exponent** and normalised to stride time (divided by stride frequency) as **Lyapunov per stride**, Lyapunov exponents are a qualitative and quantitative characterization of the dynamical behaviour of an acceleration signal [27]. | A measure of local dynamic stability of walking, which is the sensitivity of gait to small perturbations that occur naturally during walking [28].<br>*Lower values of Lyapunov estimate and Lyapunov per stride are indicative of a better gait quality.* |
| **Gait quality composite score** | Weighed sum of the root mean square in mediolateral direction, the index of harmonicity mediolateral, the magnitude of the acceleration at the dominant period in the frequency domain in the anteroposterior direction and the autocorrelation of the acceleration at the dominant period in the frequency domain [13]. | A summed gait quality score [13].<br>*A higher gait quality composite score is indicative of a better gait quality.* |

Note: <sup>a</sup> = determined in three directions: vertical, mediolateral and anteroposterior direction. psd = power spectral density.

#### 2.2.3. Socio-demographic, clinical and daily-life activity characteristics

In DIALOAD and DWELL-NL, participants were screened for the following socio-demographic and clinical characteristics: age, gender, BMI, diabetes type, employment, living situation and educational level.

In both studies, daily-life activity characteristics were derived from the tri-axial accelerometer data collected over the 7-day period (paragraph 2.2.2.). The manufacturer’s algorithm was used to identify the following daily-life activity characteristics: number of steps, number of episodes and duration for walking, standing, sitting and lying. At least four valid days were required for analysis [29]; a day was valid if the accelerometer was worn at least 12 hours in day time [30]. Daily-life activity characteristics were averaged over the valid days.

In both studies, participants performed at baseline the Short Physical Performance Battery (SPPB), a physical performance test consisting of three domains: standing balance, gait speed test and timed 5-repetition sit-to-stand test [31]. The SPPB was assessed and automatically scored using a wearable sensor solution (MoveTest, McRoberts, Den Haag, the Netherlands), containing 3D accelerometers and gyroscopes, worn dorsally at vertebra L5 using an elastic belt. Higher scores represent better physical performance (range score: 0-12).

### 2.3. Statistical analysis

Socio-demographic, clinical and daily-life activity characteristics were compared between the DIALOAD and DWELL-NL study, and between people with diabetes and peripheral neuropathy and people with diabetes without peripheral neuropathy using the Students’ t-test for continuous variables and Chi-square for categorical variables. To investigate whether peripheral neuropathy was associated with gait quality characteristics, linear regression analyses were performed. Before conducting these analyses, we checked the assumptions of linear regression. Because of the exploratory study design, we did not adjust for multiple testing [32]. The independent variable was peripheral neuropathy and the dependent variables were gait quality characteristics. Since walking speed is a determinant of many other gait quality characteristics [12,20], all analyses were adjusted for walking speed. Also, the confounding effect of the following variables was tested: age, BMI, gender and type of diabetes. If a variable changed the regression coefficient with 10% or more, this variable was considered a confounder [33]. Sample entropy in all directions was transformed by base-10 logarithm because of a non-normal distribution. We used a convenience sample to conduct this exploratory study. All statistical analysis were performed using SPSS Statistics for Windows, version 28 (IBM Corp., Armonk, N.Y., USA) with significance level of α<0.05.

## 3. Results

In DIALOAD we excluded participants aged <55 years (n=8); compared to participants aged ≥55 years (n=52), the former had more often type 1 diabetes, were more often employed, scored higher on the performance test and were more active during the day (Appendix A). Of the 52 included DIALOAD participants at baseline, we excluded one participant with insufficient raw accelerometer data and one participant with <50 walking episodes, resulting in a total of 50 included DIALOAD participants. Of the 56 included DWELL-NL participants at baseline, we excluded six participants with insufficient raw accelerometer data, one participant with <50 walking episodes, and one participant with a missing peripheral neuropathy classification, resulting in a total 48 included DWELL-NL participants.

Socio-demographic, clinical, and daily-life activity characteristics of participants in the DIALOAD and DWELL-NL studies, and people with and without peripheral neuropathy are shown in Table 2. Compared to people without peripheral neuropathy, those with neuropathy were more often male, were taller, had a different distribution in education level, had shorter daily standing time, and fewer sitting episodes. Other baseline and daily-life activity characteristics were not statistically significantly different between groups.

**Table 2.** Socio-demographic, clinical and daily-life activity characteristics.

| Socio-demographic and clinical characteristics | All participants (n=98) | DIALOAD (n=50) | DWELL-NL (n=48) | People with peripheral neuropathy (n=68) | People without peripheral neuropathy (n=30) |
| --- | --- | --- | --- | --- | --- |
| Age (years) | 68.2 (SD 7.7) | 67.7 (SD 7.3) | 68.7 (SD 8.1) | 69.0 (SD 7.8) | 66.2 (SD 7.2) |
| Female sex | 33% (n=32) | 20% (n=10) ** | 46% (n=22) ** | 24% (n=16)** | 53% (n=16)** |
| Height (m) | 176.0 (SD 11.6) | 182.3 (SD 10.5) *** | 169.4 (SD 8.8) *** | 179.3 (SD 11.6) *** | 168.7 (SD 8.0) *** |
| BMI (kg/m<sup>2</sup>) | 30.2 (SD 5.3) | 29.6 (SD 5.4) | 30.7 (SD 5.3) | 29.9 (SD 5.3) | 30.8 (SD 5.4) |
| Type 2 diabetes | 94% (n=92) | 88% (n=44) * | 100% (n=48) * | 91% (n=62) | 100% (n=30) |
| Presence of peripheral neuropathy | 69% (n=68) | 100% (n=50) | 38% (n=18) | 100% (n=68) | 0% (n=0) |
| Employed | 31% (n=30) | 34% (n=17) | 27% (n=13) | 29% (n=20) | 33% (n=10) |
| Living alone | 39% (n=38) | 48% (n=24) | 29% (n=14) | 43% (n=29) | 30% (n=9) |
| Education level |  | ** | ** | *** | *** |
| - Low | 16% (n=16) | 26% (n=13) | 6% (n=3) | 22% (n=15) | 3% (n=1) |
| - Moderate | 37% (n=36) | 22% (n=11) | 52% (n=25) | 25% (n=17) | 63% (n=19) |
| - High | 47% (n=46) | 52% (n=26) | 42% (n=20) | 53% (n=36) | 33% (n=10) |
| Short Physical Performance Battery score (range score: 0-12) | 9.0 [IQ 7.0; 10.0] (n=71) | 9.0 [IQ 7.0; 10.0] (n=38) | 9.0 [IQ 6.0; 11.0] (n=33) | 9.0 [IQ 6.0; 10.0] (n=50) | 10.0 [IQ 7.5; 11.0] (n=21) |
| **Daily-life activity characteristics** | **(n=93)** | **(n=47)** | **(n=46)** | **(n=64)** | **(n=29)** |
| Walking |  |  |  |  |  |
| - Number of steps per day | 5372 (SD 3211) | 5213 (SD 3404) | 5534 (SD 3031) | 5250 (SD 3145) | 5640 (SD 3394) |
| - Number of episodes per day | 378 (SD 175) | 349 (SD 182) | 408 (SD 164) | 362 (SD 172) | 415 (SD 179) |
| - Duration per day (hours) | 1.1 (SD 0.6) | 1.1 (SD 0.6) | 1.1 (SD 0.5) | 1.1 (SD 0.6) | 1.1 (SD 0.6) |
| Standing |  |  |  |  |  |
| - Number of episodes per day | 803 (SD 335) | 744 (SD 345) | 863 (SD 318) | 772 (SD 329) | 870 (SD 346) |
| - Duration per day (hours) | 2.2 (SD 0.9) | 1.8 (SD 0.7) *** | 2.5 (SD 1.0) *** | 2.0 (SD 0.8) ** | 2.6 (SD 1.1) ** |
| Sitting |  |  |  |  |  |
| - Number of episodes per day | 110 (SD 52) | 91 (SD 25) *** | 129 (SD 65) *** | 99 (SD 39) * | 133 (SD 68) * |
| - Duration per day (hours) | 9.5 (SD 2.5) | 9.4 (SD 2.7) | 9.5 (SD 2.3) | 9.6 (SD 2.6) | 9.7 (SD 2.3) |
| Lying |  |  |  |  |  |
| - Number of episodes per day | 12 (SD 9) | 11 (SD 5) | 14 (SD 12) | 11 (SD 7) | 15 (SD 14) |
| - Duration per day (hours) | 9.6 (SD 2.4) | 9.7 (SD 2.8) | 9.5 (SD 2.0) | 9.5 (SD 2.5) | 9.7 (SD 2.3) |
| Total wearing time per day (hours) | 22.7 (SD 2.1) | 22.3 (SD 2.4) | 23.1 (SD 1.8) | 22.6 (SD 2.1) | 22.9 (SD 2.2) |

Note: Continuous data are mean (SD standard deviation) if normally distributed and median [IQ 25th percentile; 75th percentile] if not normally distributed, and discrete data are percentage (number of). Significant differences are depicted as \*\*\* = $p < 0.001$ , \*\* = $p < 0.01$ and \* = $p < 0.05$ . If data was missing for a characteristic, the number of available data is shown in superscript.

In Table 3 the gait quality characteristics for people with and without peripheral neuropathy are shown, as well as the associations between peripheral neuropathy and gait quality characteristics, adjusted for confounders. Peripheral neuropathy was significantly associated with a lower walking speed (unstandardised β: -0.114, 95% confidence interval (CI): -0.202 to -0.026, p=0.012), a lower stride frequency (unstandardised β: -0.030, 95% CI: -0.057 to -0.003, p=0.028), lower gait intensity (i.e. lower root mean square) in vertical direction (unstandardised β: -0.074, 95% CI: -0.143 to -0.006, p=0.034), and less gait symmetry (i.e. lower harmonic ratio) in vertical direction (unstandardised β: - 0.322, 95% CI: -0.474 to -0.170, p<0.001). People with peripheral neuropathy had non-significantly poorer gait quality for most of the other 21 gait quality characteristics (Tables 1 and 3).

**Table 3.** Association between the presence of peripheral neuropathy and daily-life gait quality characteristics.

|  | Total group (n=98) | People with peripheral neuropathy (n=68) | People without peripheral neuropathy (n=30) | Total group (n=98) |  |
| --- | --- | --- | --- | --- | --- |
| | Mean (SD) | Mean (SD) | Mean (SD) | Unstandardised $\beta$ (95% CI) | p-value |
| **Walking speed (m/s)** | 0.83 (SD 0.20) | 0.81 (SD 0.19) | 0.88 (SD 0.23) | -0.114 (-0.202 to -0.026) <sup>g</sup> | 0.012* |
| **Stride length (m)** | 1.07 (SD 0.20) | 1.06 (SD 0.20) | 1.10 (SD 0.20) | 0.028 (-0.011 to 0.067) <sup>w,g,a</sup> | 0.159 |
| **Stride length variability (m)** | 0.07 (SD 0.02) | 0.07 (SD 0.02) | 0.07 (SD 0.02) | -0.008 (-0.016 to 0.000) <sup>w,g,a,d</sup> | 0.062 |
| **Stride frequency (strides/s)** | 0.82 (SD 0.07) | 0.81 (SD 0.07) | 0.85 (SD 0.07) | -0.030 (-0.057 to -0.003) <sup>w</sup> | 0.028* |
| **Stride regularity VT** | 0.60 (SD 0.13) | 0.59 (SD 0.12) | 0.64 (SD 0.13) | -0.009 (-0.045 to 0.028) <sup>w,d</sup> | 0.641 |
| **Stride regularity ML** | 0.52 (SD 0.10) | 0.52 (SD 0.11) | 0.52 (SD 0.10) | 0.029 (-0.018 to 0.077) <sup>w,g,d</sup> | 0.224 |
| **Stride regularity AP** | 0.53 (SD 0.11) | 0.53 (SD 0.11) | 0.54 (SD 0.10) | 0.037 (-0.007 to 0.082) <sup>w,d,g</sup> | 0.099 |
| **Gait complexity** |  |  |  |  |  |
| Sample entropy VT <sup>a</sup> | 0.27 (SD 0.09) | 0.27 (SD 0.06) | 0.26 (SD 0.13) | 1.023 (0.938 to 1.117) <sup>w,d,b</sup> | 0.614 |
| Sample entropy ML <sup>a</sup> | 0.31 (SD 0.09) | 0.30 (SD 0.07) | 0.32 (SD 0.12) | 0.931 (0.839 to 1.030) <sup>w,d,g,a</sup> | 0.166 |
| Sample entropy AP <sup>a</sup> | 0.27 (SD 0.09) | 0.27 (SD 0.08) | 0.28 (SD 0.12) | 0.912 (0.828 to 1.005) <sup>w,b,d,g</sup> | 0.061 |
| **Gait intensity** |  |  |  |  |  |
| Root mean square VT (m/s<sup>2</sup>) | 1.46 (SD 0.48) | 1.38 (SD 0.43) | 1.63 (SD 0.55) | -0.074 (-0.143 to -0.006) <sup>w</sup> | 0.034* |
| Root mean square ML (m/s<sup>2</sup>) | 1.23 (SD 0.26) | 1.19 (SD 0.25) | 1.31 (SD 0.27) | -0.054 (-0.151 to 0.043) <sup>w,g,a</sup> | 0.273 |
| Root mean square AP (m/s<sup>2</sup>) | 1.10 (SD 0.27) | 1.06 (SD 0.26) | 1.18 (SD 0.29) | -0.015 (-0.075 to 0.046) <sup>w,g,a</sup> | 0.631 |
| **Gait smoothness** |  |  |  |  |  |
| Index of harmonicity VT | 0.63 (SD 0.15) | 0.62 (SD 0.15) | 0.65 (SD 0.15) | 0.017 (-0.036 to 0.070) <sup>w,g,d,b</sup> | 0.523 |
| Index of harmonicity ML | 0.48 (SD 0.21) | 0.50 (SD 0.22) | 0.43 (SD 0.19) | 0.008 (-0.075 to 0.091) <sup>w,g,a</sup> | 0.847 |
| Index of harmonicity AP | 0.70 (SD 0.09) | 0.70 (SD 0.09) | 0.69 (SD 0.09) | 0.001 (-0.036 to 0.039) <sup>w,g,a</sup> | 0.937 |
| **Gait symmetry** |  |  |  |  |  |
| Harmonic ratio VT | 1.96 (SD 0.51) | 1.82 (SD 0.44) | 2.27 (SD 0.55) | -0.322 (-0.474 to -0.170) <sup>w</sup> | <0.001*** |
| Harmonic ratio ML | 1.84 (SD 0.30) | 1.80 (SD 0.28) | 1.91 (SD 0.34) | -0.034 (-0.168 to 0.101) <sup>w,d,g,a</sup> | 0.622 |
| Harmonic ratio AP | 1.77 (SD 0.42) | 1.72 (SD 0.42) | 1.87 (SD 0.40) | -0.017 (-0.163 to 0.130) <sup>w,g,d,a</sup> | 0.823 |
| **Gait consistency** |  |  |  |  |  |
| Power at step frequency VT (psd) | 0.56 (SD 0.21) | 0.53 (SD 0.19) | 0.63 (SD 0.23) | -0.016 (-0.074 to 0.042) <sup>w,g,b,d</sup> | 0.587 |
| Power at step frequency ML (psd) | 0.39 (SD 0.20) | 0.40 (SD 0.21) | 0.36 (SD 0.17) | 0.012 (-0.069 to 0.093) <sup>w,g,b,d</sup> | 0.762 |
| Power at step frequency AP (psd) | 0.54 (SD 0.14) | 0.54 (SD 0.14) | 0.54 (SD 0.13) | 0.009 (-0.055 to 0.074) <sup>w,g,a,d</sup> | 0.775 |
| **Gait stability** |  |  |  |  |  |
| Lyapunov estimate | 2.08 (SD 0.22) | 2.09 (SD 0.22) | 2.07 (SD 0.25) | -0.056 (-0.149 to 0.038) <sup>w,d,g</sup> | 0.242 |
| Lyapunov per stride | 2.57 (SD 0.33) | 2.61 (SD 0.29) | 2.48 (SD 0.38) | 0.036 (-0.072 to 0.144) <sup>w,d</sup> | 0.506 |
| **Gait quality composite score** | 0.71 (SD 0.77) | 0.64 (SD 0.80) | 0.87 (SD 0.69) | 0.082 (-0.201 to 0.365) <sup>w,d,g,a</sup> | 0.566 |

Note: <sup>a</sup> = Sample entropy in VT, ML and AP direction was transformed by base-10 logarithm for the linear regression analysis because of a non-normal distribution, and in this table the data was back-transformed for data interpretation. The shaded rows are the significant associations and these are depicted as \*\*\* = $p < 0.001$ , \*\* = $p < 0.01$ and \* = $p < 0.05$ . SD = standard deviation, CI = confidence interval, VT = vertical direction, ML = mediolateral direction, AP = anteroposterior direction, BMI = body mass index. Adjusted for the following confounders: <sup>a</sup> = age, <sup>b</sup> = BMI, <sup>d</sup> = diabetes type, <sup>g</sup> = gender and <sup>w</sup> = walking speed.

## 4. Discussion

In this study, we explored the association between peripheral neuropathy and gait quality characteristics in daily life in people with diabetes. Four of the 25 assessed daily-life gait quality characteristics were significantly lower in people with diabetes and peripheral neuropathy (i.e. walking speed, stride frequency, gait intensity and gait symmetry), indicating poorer gait quality. The other gait quality characteristics did not show significant differences, but most pointed towards poorer gait quality for people with diabetes and peripheral neuropathy. Taken together, we found in this study that peripheral neuropathy negatively affects gait quality in people with diabetes.

Peripheral neuropathy was associated with a lower walking speed and lower stride frequency, both indicating a poorer gait quality [12]. In addition, peripheral neuropathy was associated with lower gait intensity (i.e. root mean square) and less gait symmetry (i.e. harmonic ratio) in the vertical direction, indicating less trunk movement during walking and less symmetry between the left and right steps in the vertical direction, respectively [24,26]. Our results correspond with the in-laboratory findings of Menz and colleagues [34], who found poorer gait quality in people with diabetes and peripheral neuropathy compared to community-dwelling people. Furthermore, other studies found a significant association between falling and the above mentioned gait quality characteristics in community-dwelling elderly: fallers had a lower walking speed, lower stride frequency, and lower gait intensity and less gait symmetry in vertical direction than non-fallers [11,25]. Our results suggest that peripheral neuropathy leads to negative changes in daily-life gait, which may be an important factor in the increased fall risk in this population.

Three of the four significantly associated gait characteristics – walking speed, and gait intensity and gait symmetry in the vertical direction – are estimated by the vertical displacement of the trunk (i.e. upward and downward movements) [18]. The vertical accelerations are generated by the support moment of the supported leg during the gait cycle, and are the sum of ankle plantar flexion, knee extension and hip extension moments [35]. The support moment pushes the body against the ground during the stance phase of walking, and resists the lower leg from collapsing [35]. Therefore, this support moment plays an important role in maintaining balance and controlling position of the lower limb segments. It seems that the support moment is affected in people with diabetes and peripheral neuropathy, which may lead to instability in the lower leg and subsequently to a poorer gait quality.

The majority of estimated gait quality characteristics had large standard deviations, showing heterogeneity between participants. These large standard deviations were comparable to other studies that analysed the same gait quality characteristics in community-dwelling elderly [10–12]. Despite the large standard deviations, and even after adjusting for walking speed in the regression analysis, significant associations with peripheral neuropathy persisted for stride frequency, gait intensity and gait symmetry in the vertical direction. This suggests that these characteristics are all affected by peripheral neuropathy. A lower walking speed itself was also associated with peripheral neuropathy, indicating poorer gait quality [12]. Other studies found that a lower walking speed was associated with higher risk on mortality and cardiovascular disease [36]. This suggests that walking speed may be reflective of the general health of older adults, and could therefore be used as an easy-to-use biomarker [36]. This exploratory study provides a basis for future research focussing on daily-life gait analysis in people with diabetes, a topic lacking research. Considering the large standard deviations and the presence of only four statistically significant associations, we recommend to conduct this type of research in larger populations in future studies.

Compared to people with diabetes without peripheral neuropathy, those with neuropathy had a shorter standing time and fewer sitting episodes in daily life; however, there was no difference in walking activity level (i.e. daily number of steps, and walking episodes and duration). Moreover, previous studies have shown that the number of steps is not discriminative between people with diabetes and peripheral neuropathy, and those without peripheral neuropathy or even other populations [37]. This suggests that peripheral neuropathy and its negative impact on gait quality do not seem to be a cause nor a consequence of walking activity level in daily life. This highlights the importance of comprehensive gait analysis, as done in this study, for a thorough understanding of the impact that peripheral neuropathy has on daily-life gait quality in people with diabetes.

Our study has several strengths and limitations. A strength of this study was that we assessed gait quality in people with diabetes by obtaining the data in their own environment during their regular daily lives. Furthermore, we used reliable methods to estimate gait quality characteristics from raw acceleration data [9,10]. In addition, we merged two datasets (DIALOAD and DWELL-NL), which enriched the diversity of participants, and thereby increased the generalisability of this study. A limitation of our study was that we did not analyse walking episodes shorter than 5 seconds and therefore missed some walking episodes. However, these shorter walking episodes are unreliable and were therefore excluded from the analysis [38]. Second, walking speed was based on an estimated leg length of 53% of body height [19], because leg length was not individually measured in all participants. There may be some variation in the ratio of leg length to body height between individuals, however, using a standard ratio does not introduce measurement differences between researchers.

Despite the need for more confirmatory research, we believe that the current findings suggest there is potential in using gait training to improve gait quality as target for fall prevention programs in people with diabetes and peripheral neuropathy. A systematic review found that a combination of gait, balance, and functional training did not reduce the risk of falling in people with peripheral neuropathy [39]; however, a meta-analysis conducted in community-dwelling adults demonstrated its effectiveness [40]. This suggests that the existing gait-training methods are rather general and may not be applicable to people with peripheral neuropathy – who have an elevated fall risk [5]. This highlights the need to develop specific gait training methods tailored to address the gait quality characteristics affected in people with peripheral neuropathy.

To conclude, in this exploratory study, we found that people with diabetes and peripheral neuropathy had a lower walking speed, lower stride frequency, lower gait intensity, and less gait symmetry than their peers with diabetes but without peripheral neuropathy, indicating poorer gait quality. Other parameters, although not significantly associated, pointed in the same direction of poorer gait quality in people with peripheral neuropathy. These results may help to develop targeted gait training to improve gait quality and potentially reduce fall risk in people with peripheral neuropathy.

## Data Availability

All data produced in the present study are available upon reasonable request to the authors

## Author contributions: Chantal M Hulshof

Conceptualization, Methodology, Software, Validation, Formal analysis, Investigation, Resources, Data curation, Writing – Original Draft, Visualization, Project administration. **Marike van der Leeden:** Conceptualization, Methodology, Validation, Formal analysis, Writing – Review & Editing, Supervision, Funding acquisition. **Jaap J van Netten:** Conceptualization, Methodology, Validation, Investigation, Resources, Data curation, Writing – Review & Editing, Supervision, Project administration, Funding acquisition. **Maarten Gijssel:** Investigation, Resources, Data curation, Writing – Review & Editing, Project administration, Funding acquisition. **Jordi Evers:** Resources, Data curation, Writing – Review & Editing, Funding acquisition. **Sicco A Bus:** Conceptualization, Writing – Review & Editing, Supervision, Funding acquisition. **Mirjam Pijnappels:** Conceptualization, Software, Writing – Review & Editing, Supervision, Funding acquisition.

## Declarations of interest

None.

## Acknowledgements

The DIALOAD project was supported by the Amsterdam Movement Sciences research institute and ZGT Wetenschapsfonds. The DWELL-NL project has received funding from the Interreg 2 Seas programme 2014-2020 co-funded by the European Regional Development Fund under subsidy contract No 2S01-058.

We thank Richard Casius (Vrije Universiteit Amsterdam) for his assistance with using the Gait Analysis Toolbox, Abe Funnekotter and Renée Kers (both, Vrije Universiteit Amsterdam) for testing the Gait Analysis Toolbox on the DIALOAD data, and Marleen Eising-de Vries (Move2be) and Natasja de Graaf-Hilverts (BuurtFysio) for collecting the DWELL-NL data.

## Notes

### Competing Interest Statement

The authors have declared no competing interest.

### Author Declarations

Ethics committees of Amsterdam UMC and Brabant waived ethical approval for this work.

