## Appendix A for "The association between peripheral neuropathy and daily-life gait quality characteristics in people with diabetes"

*Table A1 – Socio-demographic, clinical and daily-life activity characteristics of DIALOAD participants aged ≥55 and <55 years*

| **Socio-demographic and clinical characteristics** | **DIALOAD ≥55 years (n=50)** | **DIALOAD <55 years (n=8)** | **p-value** |
|---|---|---|---|
| Age (years) | 67.7 (SD 7.3) | 49.8 (SD 3.7) | <0.001*** |
| Female sex | 20 (10) | 13 (1) | 0.615 |
| Height (cm) | 182.3 (SD 10.5) | 182.4 (SD 5.0) | 0.993 |
| BMI (kg/m$^2$) | 29.6 (SD 5.4) | 30.2 (SD 6.3) | 0.809 |
| Type 2 diabetes | 88 (44) | 38 (3) | <0.001*** |
| Presence of peripheral neuropathy | 100 (50) | 100 (8) | 1.000 |
| Employed | 34 (17) | 100 (8) | <0.001*** |
| Living alone | 48 (24) | 25 (2) | 0.225 |
| Education level | | | 0.209 |
| - Low | 26 (13) | 25 (2) | |
| - Moderate | 22 (11) | 50 (4) | |
| - High | 52 (26) | 25 (2) | |
| Short Physical Performance Battery score (0-12) | 9.0 [IQ 7.0; 10.0] $^{(n=38)}$ | 12.0 [IQ 11.0; 12.0] $^{(n=6)}$ | <0.001*** |
| **Daily-life activity characteristics** | **(n=47)** | **(n=8)** | |
| Walking | | | |
| - Number of steps per day | 5213 (SD 3404) | 9292 (SD 3165) | 0.003** |
| - Number of episodes per day | 349 (SD 182) | 560 (SD 561) | 0.006** |
| - Duration per day (hours) | 1.1 (SD 0.6) | 1.8 (SD 0.6) | 0.002** |
| Standing | | | |
| - Number of episodes per day | 744 (SD 345) | 1201 (SD 501) | 0.002** |
| - Duration per day (hours) | 1.8 (SD 0.7) | 2.9 (SD 0.8) | <0.001*** |
| Sitting | | | |
| - Number of episodes per day | 91 (SD 25) | 135 (SD 73) | 0.127 |
| - Duration per day (hours) | 9.4 (SD 2.7) | 7.3 (SD 1.5) | 0.037* |
| Lying | | | |
| - Number of episodes per day | 11 (SD 5) | 12 (SD 7) | 0.679 |
| - Duration per day (hours) | 9.7 (SD 2.8) | 8.8 (SD 3.4) | 0.384 |
| Total wearing time per day (hours) | 22.3 (SD 2.4) | 21.4 (SD 3.5) | 0.295 |

*Note: Continuous data are mean (SD standard deviation) if normally distributed and median [IQ 25th percentile; 75$^{th}$ percentile] if not normally distributed, and discrete data are percentage (number of). Significant differences are depicted as *** = p < 0.001, ** = p < 0.01 and * = p < 0.05. If data was missing for a characteristic, the number of available data is shown in superscript.*
